# The diagnostic accuracy of chest Xray screening for silicosis: A systematic review, meta-analysis and modelling study

**DOI:** 10.1101/2025.05.28.25328086

**Authors:** Patrick Howlett, Ashwin Durairaj, Maia Lesosky, Johanna Feary

## Abstract

**Objectives:** Chest Xray (CXR) is widely used for silicosis diagnosis, despite concerns regarding sensitivity. We investigated the diagnostic accuracy of CXR for silicosis screening compared to computed tomography (CT), high-resolution CT (HRCT) and autopsy, and modelled the relationship between CXR sensitivity and disease severity.

**Methods:** Medline, Embase, Scopus, and Web of Science databases were searched on 2^nd^ July 2024 (Prospero registration: CRD42024513830). Meta-analyses were performed by reference standard and at increasing reference test severity cut-offs. The Quality Assessment of Diagnostic Accuracy Studies-2 (QUADAS-2) tool assessed risk of bias. In scenarios of fixed and relative sensitivity, according to disease severity, we estimated missed silicosis cases and the number needed to screen (NNS) in hypothetical populations of low (5%), medium (15%) and high (30%) silicosis prevalence.

**Results:** Twenty studies included 2156 participants and 1105 silicosis cases. CXR had moderate sensitivity (0.76; 95% confidence interval (CI): 0.63-0.86, I^2^=84%) and high specificity (0.89, 95% CI: 0.77-0.95, I^2^=57%) compared to HRCT in 12 studies, and low sensitivity (0.50, 95% CI: 0.45-0.55, I^2^=0%) and high specificity (0.91, 95% CI: 0.87-0.93, I^2^=20%) compared to autopsy in two studies. CXR sensitivity increased with higher reference test severity cut-offs. Clinically relevant numbers of cases were missed in fixed and relative sensitivity scenarios; increased prevalence and less severe disease resulted in more missed cases and a lower NNS.

**Conclusions:** Silicosis severity and reference test type both plausibly influence CXR sensitivity. Assuming either fixed or relative sensitivity results in missed silicosis cases. Judicious HRCT screening is likely to improve case detection.

**What is already known on this topic:** It is widely understood that Chest Xray (CXR) underdiagnoses silicosis compared to more accurate methods, such as high resolution computed tomography (HRCT) and autopsy.

**What this study adds:** Our systematic review and meta-analysis demonstrated that the sensitivity of CXR was lowest when compared to autopsy (50%), followed by HRCT (76%). This difference may be explained by the increased accuracy of autopsy as a reference test. Another potential explanation for differences between study results could be that – because severe silicosis is more easily diagnosed by CXR – studies with a higher proportion of severe disease recorded higher sensitivity results. Importantly, regardless of whether differences between studies are explained by different reference test modalities or the proportion of severe disease, when modelled among a population of silica-exposed workers, many silicosis cases are missed.

**How this study might affect research, practice or policy:** This study suggests the careful implementation of HRCT screening for silicosis would improve case detection.

## Introduction

Silicosis is a preventable yet incurable occupational fibrotic lung disease caused by the inhalation of respirable crystalline silica (RCS) dust. Recent reports of the high morbidity and mortality of artificial stone silicosis are reminders of the potentially devastating effects of this disease[1,2]. The likely underdiagnosis of silicosis and the lack of clarity around modelling methods mean the global burden of disease is unknown and likely higher than currently reported estimates[3,4].

Silicosis diagnosis relies on positive Chest Xray (CXR) and/or Computed Tomography (CT) findings in conjunction with a compatible exposure history. The International Labour Organisation (ILO) classification system[5] is one system that provides a standardised approach to screening for possible silicosis cases in at-risk populations. The 12-point, stepwise classification describes the severity and profusion of small-opacities, along with other relevant findings. In UK and US practice, a profusion cut off of ≥ 1/0 often qualifies as a positive screening result[6,7]; CT is normally reserved for cases with an abnormal CXR. Due to availability and cost, for most workers globally, CXR is the only test performed to diagnose silicosis. In practice, therefore, a positive screening CXR in an exposed worker will be classified as a case of silicosis. In many settings, the expertise required to apply the ILO classification precludes its use, relying instead on a simple radiographic diagnosis.

The concern that some silicosis may not be visible on CXR, sometimes termed “subradiological silicosis”, dates back to autopsy studies of South African miners, which demonstrated a sensitivity of 31% at a CXR >= 1/1 ILO cut-off[8,9]. More recently, among artificial stone workers in Australia, a CXR sensitivity of 48% was reported compared to HRCT[10]. These enduring concerns are changing practice. A 2019 Royal Australian and New Zealand College of Radiologists Position Statement strongly recommended CT scanning as the “primary imaging modality for screening exposed workers”. However, the potential limitations of CXR have not been systematically reviewed or quantified[11]. In practice, and for the purposes of this paper, HRCT and reconstructed multidetector CT (MDCT) images are collectively referred to as HRCT.

It is intuitive that CXR sensitivity is related – at least in part – to severity of disease, such that CXRs with a greater severity of disease have a higher sensitivity. In South African miners, CXR sensitivity increased from 38% in the subgroup with “slight” disease at autopsy, to 83% in those with “marked” disease[9]. The counterfactual assumes that sensitivity does not vary with silicosis severity. These two relationships are illustrated by directed acyclic diagrams in Figure 1A and 1B.

**Figure 1.**
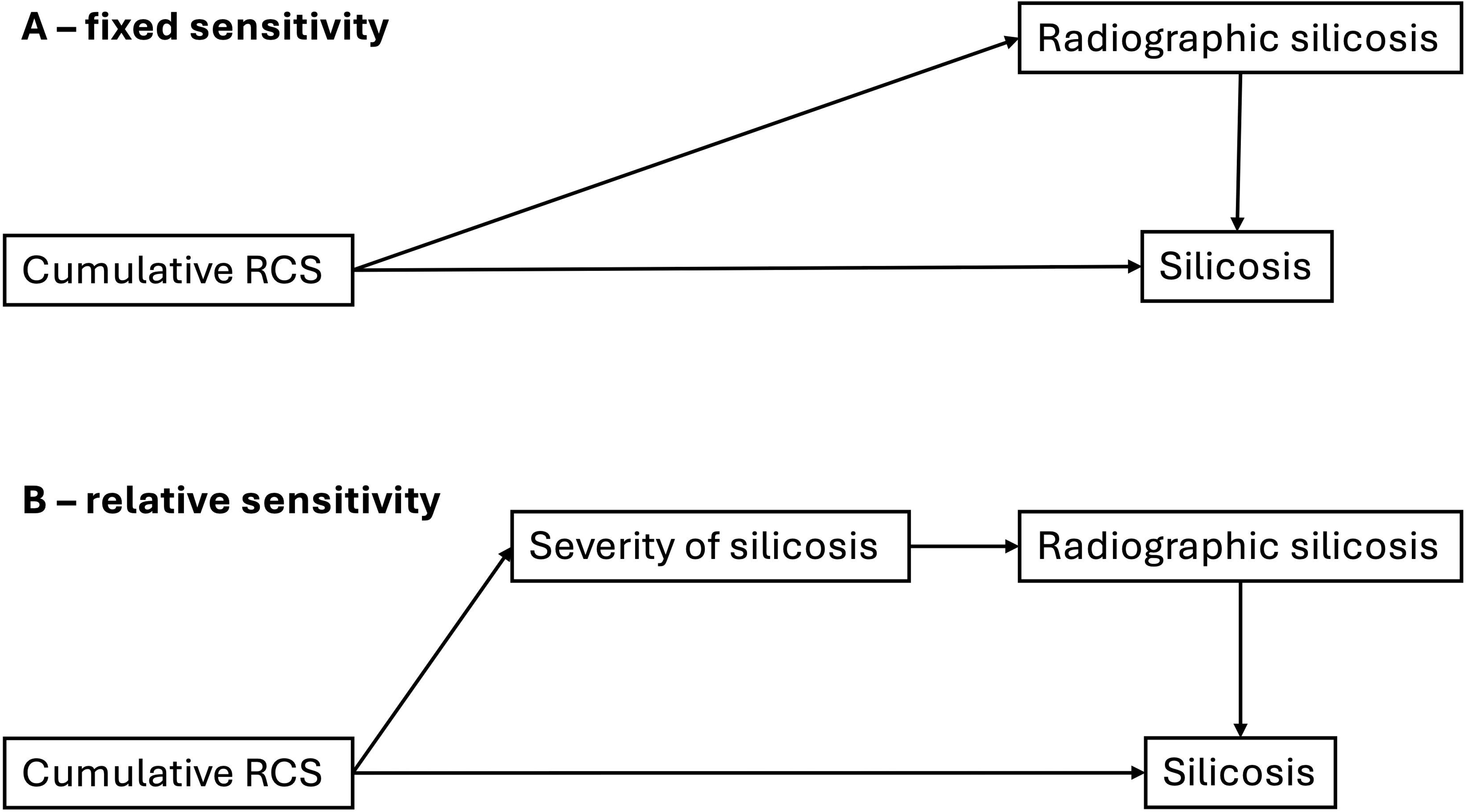
Directed acyclic diagrams of the conceptual relationship between cumulative silica exposure and silicosis. Two counterfactual relationships are represented. A. The relationship between cumulative silica exposure and radiological silicosis is fixed – sensitivity does not vary according to disease severity. B. The relationship between cumulative silica exposure and identification on radiological test is mediated and thus relative to the severity of silicosis – sensitivity varies relative to disease severity.

Quantifying the sensitivity of CXR compared to high-resolution computed tomography (HRCT) informs the debate on the role of HRCT in silicosis screening[12]. Understanding whether silicosis severity may modify CXR sensitivity is helpful clinically and programmatically. Understanding the impact of the counterfactual scenarios of fixed and relative sensitivity, according to silicosis severity, on the number missed cases may be helpful in identifying priority populations for HRCT screening.

We therefore performed a systematic review and meta-analysis of the sensitivity and specificity of CXR compared to computed tomography (CT), HRCT and autopsy reference standards. We then modelled how sensitivity and specificity of CXR changes according to severity of underlying disease. We used this model to estimate the number of cases missed and number needed to screen (NNS) in example populations of silica-exposed individuals.

## Methods

One reviewer (AD) searched Medline, Embase, Scopus and Web of Science databases from inception up to 2^nd^ July 2024 (search strategy in online supplementary materials 1) and reviewed bibliographies of included studies (Prospero ID: CRD42024513830).

Study inclusion criteria were: (1) cohort, cross-sectional, or case-control study design; (2) Population ≥18 years-old with occupational RCS exposure; (3) CXR used as the index test for assessing silicosis, using the ILO classification or equivalent; (4) CT, HRCT, or autopsy used as the reference standard for assessing silicosis; (5) sufficient information to construct 2×2 tables. In the case of a high risk of duplicated data, the most comprehensive study was retained.

Title and abstract screening and full-text review was performed in parallel by two reviewers (AD, PH) using Covidence. Data was extracted by a single reviewer (AD) and then checked (PH). Disagreements were resolved through consensus. Methodological quality was assessed using the Quality Assessment of Diagnostic Accuracy Studies-2 (QUADAS-2) tool[13] and entered into RevMan. Extraction tools are presented in online supplementary materials 2 and description of our application of the QUADAS-2 tool in supplementary materials 3.

## Meta-analysis

Our pre-specified CXR meta-analysis of sensitivity and specificity of CXR for silicosis included studies using the ILO CXR cut-off of ≥1/0 or equivalent for the index test[14]. The HRCT group included reconstructed MDCT scans, all of which were labelled as HRCT in their respective studies. Post-hoc sensitivity analyses included studies with other CXR cut-offs and restriction to studies without a high/unclear risk of patient selection or index test bias.

To demonstrate whether CXR diagnostic accuracy varied according to silicosis severity, we repeated our meta-analyses at increasing reference test severity cut-offs. We separated studies by radiology (HRCT and CT) and autopsy reference standards. Although recent consensus classifications are available[15], no single HRCT/CT or autopsy classification was consistently applied. We therefore considered four of seven studies that used a 4-level scale (similar to the ILO CXR system) as equivalent and mapped the remainder to that scale (details in online supplement materials 4).

A bivariate generalized linear mixed model (GLMM) with logit transformation was used to calculate pooled sensitivity and specificity estimates using the “meta” package in R (Version 4.3.2). The bivariate GLMM approach accounts for the correlation between sensitivity and specificity and is preferred when 2×2 cell counts are low[16]. Heterogeneity was assessed using the I^2^ statistic.

## Measures of impact

We performed a mixed-effects linear meta-regression with the exposure of the ratio of ILO category ≥2/1 relative to ≥1/0 silicosis and the outcome of CXR ≥1/0 sensitivity compared to HRCT. ILO category ≥2/1 was used as it represents major category 2 disease and provides a robust numerator. Using this model, and to demonstrate the impact of both fixed and relative sensitivity scenarios, we estimated the sensitivity of CXR for silicosis among hypothetical populations of 1000 silica-exposed workers with low (5%), medium (15%) and high (30%) prevalences of CXR ILO ≥1/0 category silicosis. We also assumed two fixed proportions of less severe disease (20% CXR ILO ≥2/1 relative to ILO ≥1/0 silicosis) and more severe disease (40% CXR ILO ≥2/1 relative to ILO ≥1/0 silicosis). In each population, we then calculated the number of missed cases that would be identified by HRCT and the number needed to screen using HRCT to detect a single extra case of silicosis. A sensitivity analysis used the prevalence of CXR (non-reference confirmed) CXR ILO category ≥2/1 silicosis as the meta-regression exposure.

All code and data are publicly available at https://github.com/pjhowlett/da_silic_cxr/tree/main.

Human Research Ethics Committee approval was not required as only previously published data was used.

## Results

From a total of 825 title and abstracts, 47 studies underwent full-text screening and 20 were included (Figure 2). Fifteen studies used HRCT, three used CT and two used autopsy as a reference standard. A total of 2156 participants (HRCT 1207, CT 151, Autopsy 798) were included, of whom 1105 (HRCT 587, CT 112, Autopsy 406) were identified by the reference test to have silicosis. There was at least moderate agreement for title and abstract and full-text reviews; Cohen’s Kappa 0.65 and 0.45, respectively.

**Figure 2.**
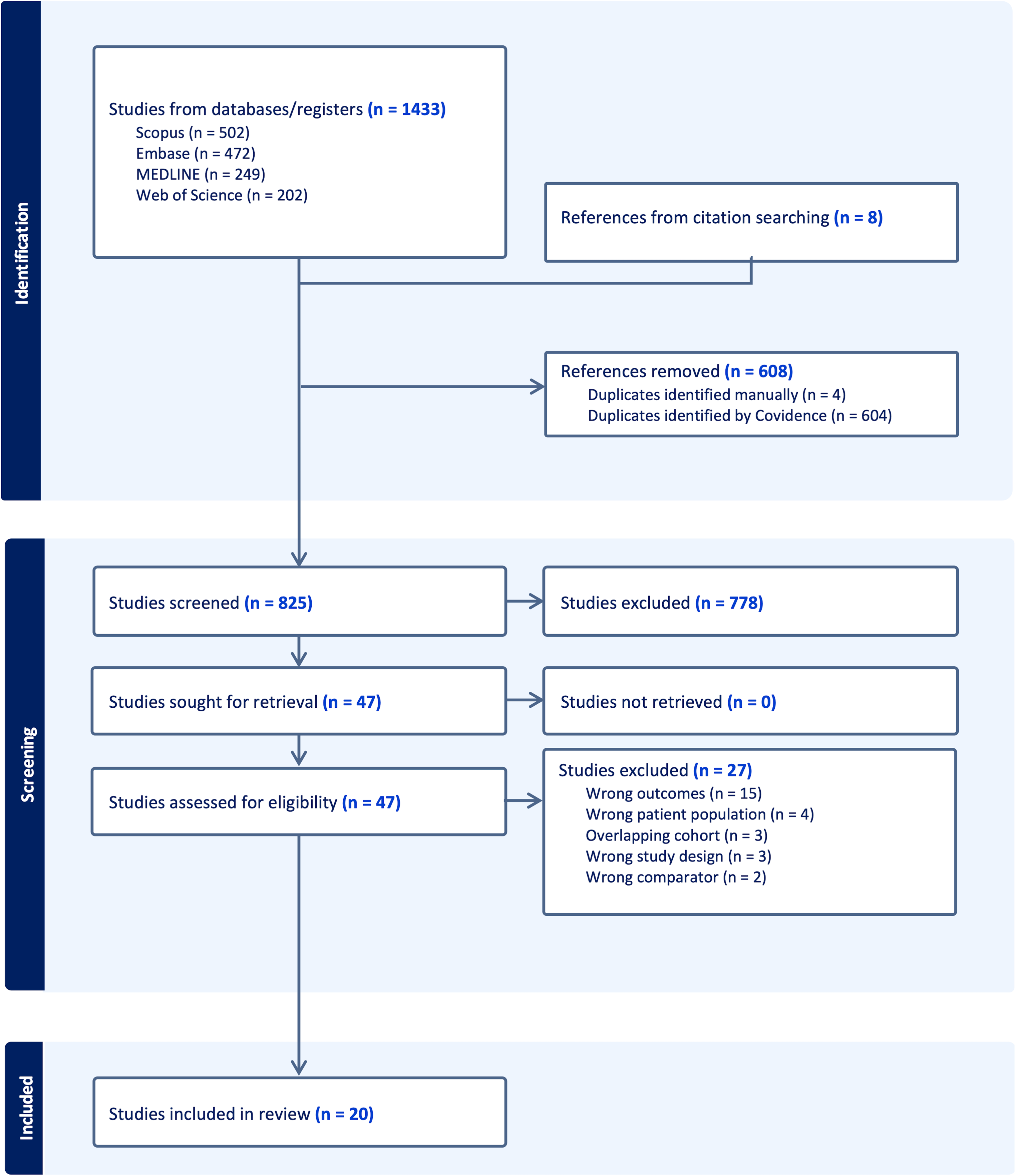
PRISMA Diagram.

All studies represented mining and non-mining industries in middle or high-income economies. The median prevalence of reference test silicosis was 63% (range 16-99%) [17–24,15,25–32,8,33,34] (online supplementary table 1). Study size ranged from 11 to 557 participants. Exposures ranged from 7.6 to 31 years; dental technician, sandblaster and artificial stone worker studies had median exposures of <15 years[25,28–30]. Autopsy and CT studies were either performed or published before HRCT in most cases, although HRCT publication dates ranged from 1991 to 2024.

Among 15 studies with HRCT as the reference test, CXR sensitivity ranged from 0.18 (95% CI 0.02, 0.52) to 0.95 (95% CI 0.76, 1.00) while specificity ranged from 0.29 (95% CI 0.04, 0.71) to 1.00 (95% CI 0.99, 1.00) (Figure 3A). For two studies with autopsy as the reference test, CXR sensitivity was 0.50 (95% CI 0.39, 0.61) and 0.50 (95%CI 0.44, 0.56), while specificity was 0.89 (95% CI 0.84, 0.93) and 0.93 (95% CI 0.87, 0.96) (Figure 3B). For three studies with CT as the reference test, CXR sensitivity ranged from 0.94 (95% CI 0.73, 1.00) to 1.00 (95% CI 0.92, 1.00) while specificity ranged from 0.50 (95% CI 0.21, 0.79) to 1.00 (95% CI 0.48, 1.00) (supplementary figure 1).

**Figure 3.**
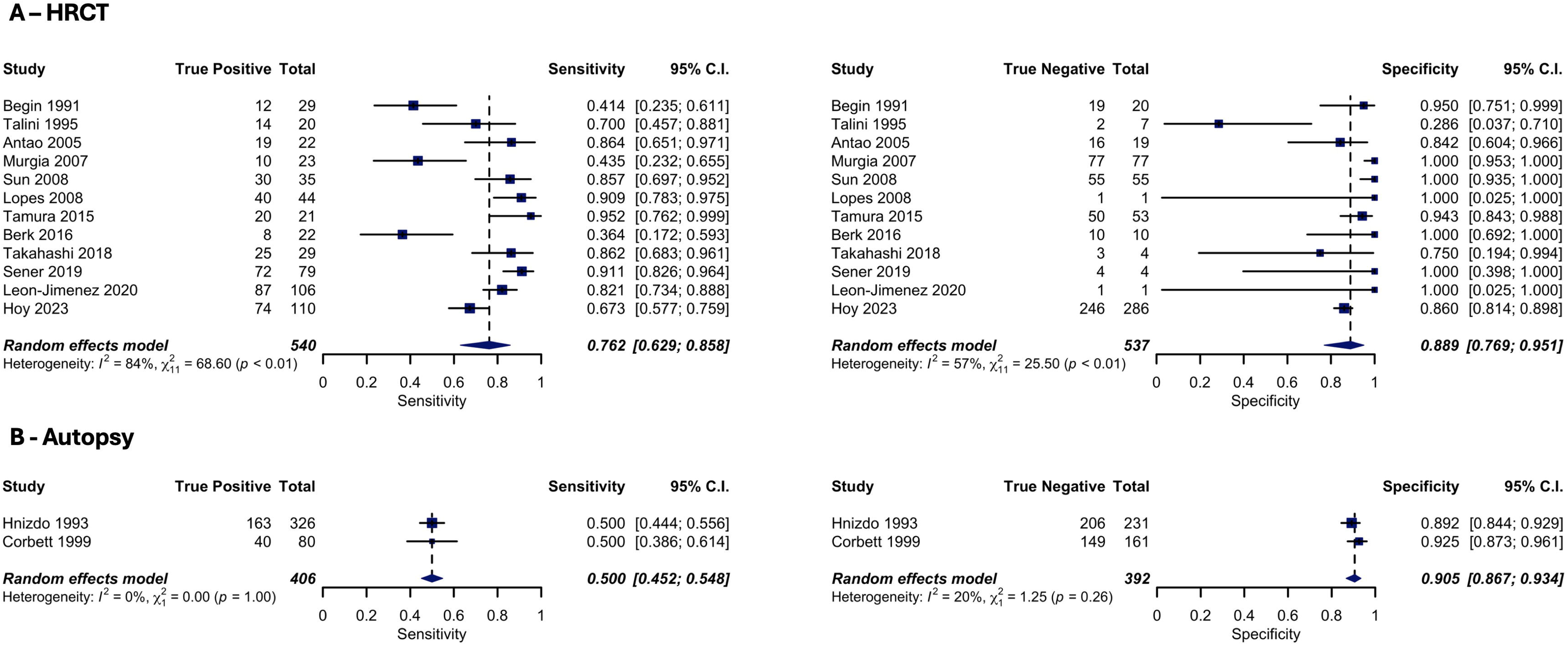
Forest plots of sensitivity and specificity from meta-analysis of chest X-ray (CXR), at an ILO >1/0 cut-off, for the diagnosis of silicosis. **A.** Describes the sensitivity (left) and specificity (right) of CXR compared to high-resolution computed tomography (HRCT). **B.** Describes the sensitivity (left) and specificity (right) of CXR compared to autopsy Random effects model uses a generalised linear mixed model. Heterogeneity is determined through the I2 statistic and Chi-squared (χ²) test. Abbreviations: C.I, confidence interval.

## Quality Assessment

Eight studies had unclear bias and three had high risk of bias regarding patient selection (online supplementary figure 2). Unclear or high risk of bias in the index (6 studies) or reference (4 studies) test was most commonly due to poor detailing of test blinding. Six studies had unclear bias and three had high risk of bias for flow and timing, due to flow concerns (2 studies) or unknown or inappropriate intervals between the index and reference test (7 studies). Five studies had high or unclear applicability, mainly related to different ILO CXR cut-offs.

## Meta-analysis

Pooled sensitivity for CXR with a HRCT reference standard was 0.76 (95% CI 0.63, 0.86; n = 12), with a pooled specificity of 0.89 (95% CI 0.77, 0.95; n = 12) (Figure 3A). Heterogeneity was substantial for sensitivity (I^2^ = 84%) and moderate for specificity (I^2^ = 57%). Compared to an autopsy reference standard, pooled sensitivity and specificity for CXR were 0.50 (95% CI 0.45, 0.55; n = 2) and 0.91 (95% CI 0.87, 0.93; n = 2), with low heterogeneity (I^2^ = 0% and I^2^ = 20%, respectively) (Figure 3B). Compared to a CT reference standard, pooled sensitivity and specificity for CXR were 0.97 (95% CI 0.86, 0.99; n = 2) and 0.69 (95% CI 0.20, 0.95; n = 2), with low (I^2^ = 0%) and moderate (I^2^ = 57%) heterogeneity, respectively (supplementary figure 1). The sensitivity analyses, which included different ILO cut-offs or the removal of studies with a high/unclear risk of patient selection bias or index test bias, did not significantly alter our results (online supplementary figures 3 and 4).

CXR demonstrated increased sensitivity at higher grade cut-offs of silicosis in the reference test (online supplementary figures 5 and 6). Sensitivity was 0.86 (95% CI 0.72, 0.93; I^2^ = 47%; n = 4) compared to combined HRCT and CT test category 1 disease or greater. All participants bar one with reference level 2 or higher disease on CT or HRCT were identified correctly by CXR. For autopsy reference standards, sensitivity increased from 0.72 (95% CI 0.27, 0.95; I^2^ = 89%; n = 3) at the lowest cut-off to 0.83 (95% CI 0.70, 0.91; I^2^ = 0%; n = 3) at the highest. Correspondingly, specificity showed a decreasing trend across both radiological and autopsy reference standards with increasing grade of cut off.

## Missed cases and number needed to screen

Our meta-regression estimated a 1.15% (95% CI 0.44, 1.86) increase in sensitivity for each 1% increase in the ratio of ILO category ≥2/1 relative to ≥1/0 silicosis. When there were no ILO category ≥2/1 silicosis cases (i.e. the ratio was zero) the sensitivity of CXR was 16%. The R^2^ value was 63% and heterogeneity substantial (I^2^ = 86%) (Figure 4).

**Figure 4.**
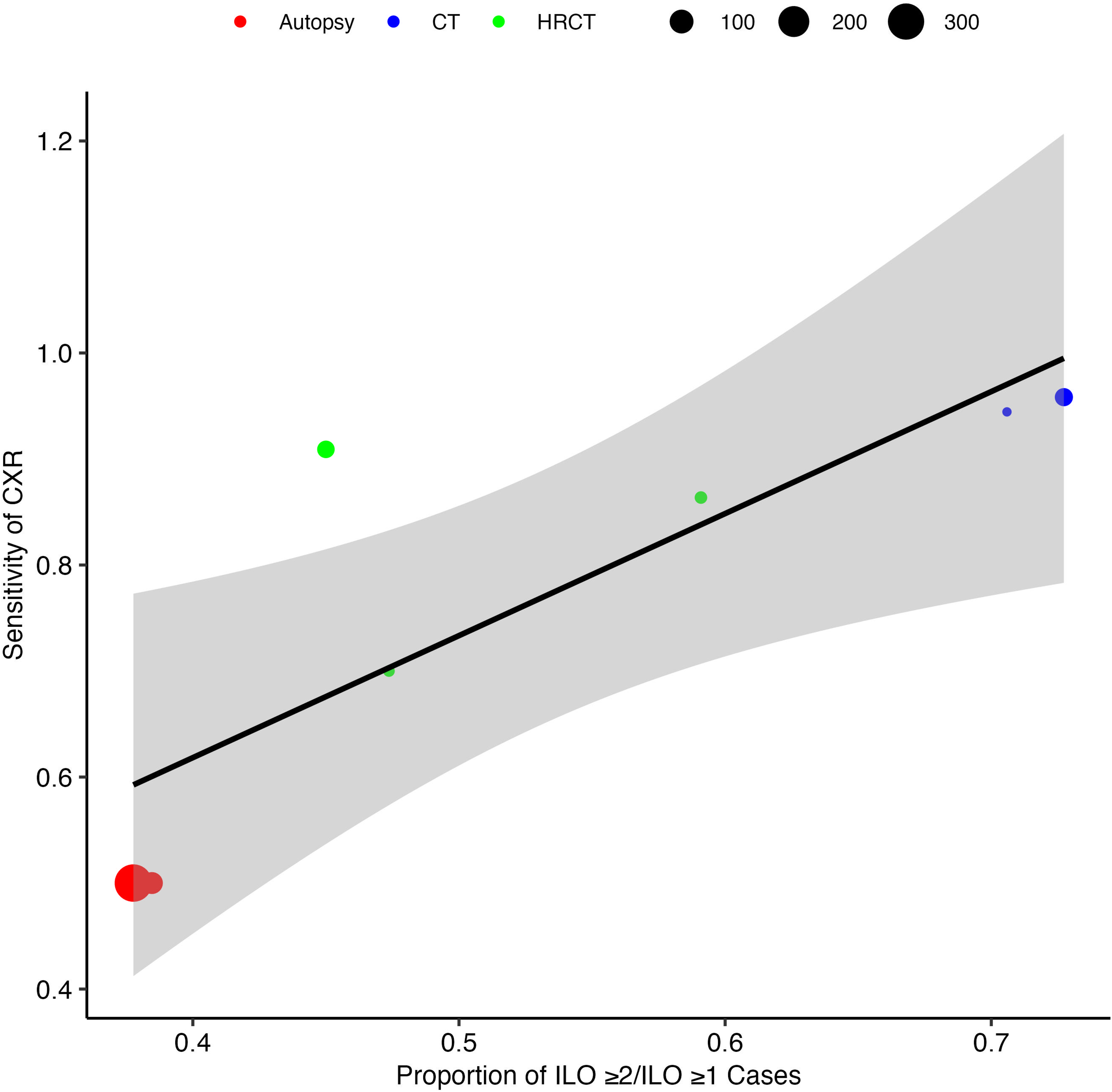
A mixed-effects linear meta-regression model showing the association between the sensitivity of CXR at ILO category ≥1 and the ratio ILO >2/1 relative to ≥1/0 silicosis across 7 studies. Each circle represents a study, with the diameter proportional to the study’s size. The circle is coloured depending on reference standard (Red = autopsy, Blue = computed tomography and Green = high resolution computed tomography). The solid line represents the predicted sensitivity based on the ratio ILO >2/1 relative to ≥1/0 silicosis. The model explains a moderate proportion of the heterogeneity in CXR sensitivity between studies (R² = 63%, I^2^ = 86%).

In a low prevalence scenario (5% or 50 silicosis cases per 1000 exposed) (Table 1 and supplementary figure 7A) and under the assumption of fixed CXR sensitivity, the number of missed cases per 1000 exposed was 16 cases and the NNS was 62 persons. Under the assumption of relative sensitivity, the number of cases missed was higher and the NNS lower; when we assumed a lower proportion of severe disease (20% cases ≥ ILO 2/1 relative to ≥1/0 silicosis), 79 cases per 1000 exposed were missed with NNS of 13 persons. When we assumed severe disease was more common (40% cases ≥ ILO 2/1 relative to ≥1/0 silicosis) 31 cases per 1000 person were missed with a NNS of 32 persons. Both increasing prevalence of silicosis and a higher proportion of severe disease led to a higher number of missed cases and, hence, a lower NNS. Our sensitivity analysis, using the prevalence of CXR ILO ≥2/1, resulted in similar estimates of missed cases and NNS (supplementary table 2, supplementary figures 7B and 8).

**Table 1.**
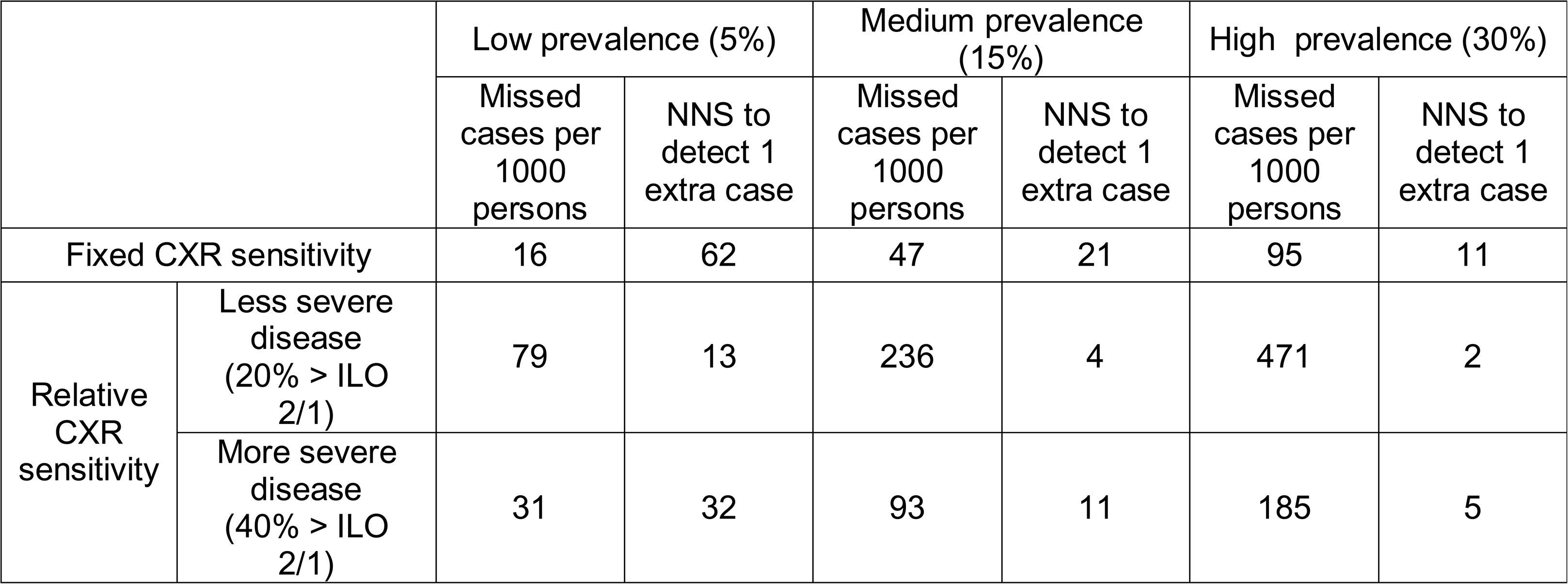
Missed cases and number needed to screen in a silica-exposed population. Calculated using the metaregression model with exposure of the ratio ILO >2/1 relative to ≥1/0 silicosis. The number of missed cases per 1000 exposed persons and number needed to screen to identify 1 extra case of silicosis in three scenarios of low silicosis prevalence (5% or 50 cases per 1000), as defined by CXR ILO >1/0, medium silicosis prevalence (15% or 150 cases per 1000) and high silicosis prevalence (30% or 300 cases per 1000), and under the assumptions of fixed and relative sensitivity of CXR, compared to HRCT. For the scenario of relative sensitivity, we have further assumed either less common severe disease (20% of cases are ≥ ILO 2/1 relative to ≥1/0 silicosis) and more common severe disease (40% of cases are ≥ ILO 2/1 relative to ≥1/0 silicosis).

## Discussion

Despite concerns regarding the diagnostic accuracy of CXR for silicosis, no systematic review or meta-analysis has been performed to date. We found that, for the diagnosis of ≥1/0 silicosis, CXR had a moderate pooled sensitivity compared to HRCT (0.76; 95% CI 0.63, 0.86) and poor sensitivity compared to autopsy (0.50; 95% CI 0.45, 0.55). Specificity was high for both autopsy and HRCT. Heterogeneity was high for HRCT (I^2^ = 84%) while two large autopsy studies reported identical sensitivity values.

The sensitivity of CXR in detecting silicosis increased across autopsy, HRCT and CT, reflecting the improved resolution and hence diagnostic accuracy of autopsy over HRCT, and HRCT over CT. That the sensitivity of CXR compared to CT approached 1 suggests that plain CT, as applied at the time of the included studies (1986-1993), had little benefit for the screening detection of silicosis beyond standard CXR. Older plain CT technology has been superseded by HRCT or reconstructed MDCT images, thus limiting the generalisability of our CT results to current CT imaging. Only one, small study from 1991 directly compared CT and HRCT; although this reported equal sensitivity, the authors commented on improved clarity and firmer interpretation with HRCT compared to CT [17]. Technological improvements over time in both CXR and reference tests may influence results. Older HRCT studies tended to have reduced sensitivity compared to more recent studies (Figure 3). This suggests greater improvement in CXR accuracy over time relative to HRCT, although these studies were also often smaller.

We observed increasing CXR sensitivity at higher cut-off grades of reference test silicosis. This supports the intuitive and previously reported understanding that severe silicosis is more readily diagnosed on CXR[8,9]. We then found that CXR sensitivity increased by 1.15% (95% CI 0.44, 1.86) for every 1% increase in the ratio of CXR ILO ≥2/1 relative to ≥1/0 silicosis, although heterogeneity was considerable (I^2^ = 86%).

Based on our findings, it is plausible that both silicosis severity and reference test type influence the diagnostic accuracy of CXR for silicosis. In the case of HRCT, it may therefore be considered that the true sensitivity and specificity of CXR lies between the assumptions of fixed and relative sensitivity. It is therefore important to note that, regardless of assumption, a clinically meaningful number of cases were missed (Table 1 and supplementary figure 7). Furthermore, higher silicosis prevalence and greater proportions of severe disease led to higher numbers of missed cases and a lower NNS. This suggests HRCT may have the greatest impact among high prevalence populations with more severe disease. Conversely, although the absolute impact of HRCT may be reduced among lower prevalence groups, if the relative sensitivity scenario is considered probable, the proportional benefit from the earlier diagnosis of silicosis is greater, particularly in the plausible case of less frequent severe disease.

From a pragmatic perspective, our results would suggest a low threshold for HRCT screening among groups or individuals considered to be at a higher risk of silicosis. From the patient and physician perspective, although no established treatment for silicosis exists, the use of HRCT may allow earlier diagnosis of silicosis, leading to more informed work choices. For employers, industrial hygienists and regulators, more accurate, earlier-stage silicosis prevalence data may prompt pro-active exposure testing, adjustments to work practices and legislation.

Given the dose-response relationship between silica and lung cancer, and often high rates of smoking among silica-exposed workers[3,35], our analysis represents the minimum potential benefit of HRCT screening. Practical considerations of HRCT screening include accessibility, cost-effectiveness, radiation dose and risks of subsequent investigations, such as lung biopsy of high-risk nodules. Correctly risk stratifying nodules among silica-exposed populations, particularly if TB is common, presents an unexplored challenge. Low-dose CT screening with high resolution reconstruction, such as implemented for lung cancer screening and silicosis screening in Western Australia, is possible[11]. An alternative approach may involve a combination of computer-aided detection for CXR, trained with HRCT as a reference standard. If successful, this approach would have global impacts for the diagnosis and classification of silicosis; our results can help design studies.

Individual study methods may influence sensitivity or specificity. Recruitment from largely pre-screened silicosis populations, in whom prevalences of silicosis are notably high (range 16-99%), may falsely raise sensitivity and reduce specificity. Differences in test reader experience or reading and consensus methods may increase between study variance. However, restriction to studies without high or unclear risk of bias in these domains did not alter our findings. Future studies should aim for an unselected population and a standardised reading and consensus approach. Comorbidities including emphysema or tuberculosis did not appear to reduce sensitivity or specificity[32,33].

Important limitations of our methods exist. Whilst we applied our risk of bias tool according to the guidance[13] and have attempted to do so transparently (including publication of our raw data), we felt the QUADAS-2 may over-estimate risk of bias in some cases. For example, the preselection of a sample (such as in Hoy et al (29)) results in a penalisation of both Patient selection and Flow and Timing, possibly over-stating the true risk of bias. Potentially exacerbated by our English-language restriction, our data is skewed to developed economies with relatively low exposures[3]. When no ILO ≥ 2/1 cases were present modelled sensitivity was 16%. This may represent inaccuracy outside our data range as our lowest ILO ≥2/1 relative to ≥1/0 percentage was 38%. However, one study of only ILO 0 and 0/1 CXRs in miners found 57/339 (17%) had silicosis on HRCT suggesting this low sensitivity is plausible[36]. Wide confidence intervals in our meta-regression represent data sparsity and heterogeneity and lend caution to our subsequent missed cases and NNS calculations. Whilst we maintain these estimates are useful, they should be viewed as illustrative estimates of the impact of changes in parameters – such as proportion of severe disease – rather than exact figures. Our sensitivity analysis (using coefficients from the prevalence of ILO ≥2/1 exposure model) demonstrates similar values, particularly at higher prevalences of silicosis. At lower ranges, the differences between the missed case and NNS estimates of our primary and sensitivity analysis suggests further caution regarding figures outside of our data range. Whilst severity of disease is likely a major determinant of sensitivity, other important factors that we did investigate that may determine CXR sensitivity include the quality of images and experience of the reader and who hired them [37].

Compared to HRCT, CXR has moderate sensitivity and high specificity for silicosis diagnosis. Both the type of reference test and the proportion of severe disease plausibly influence CXR sensitivity. Regardless, using CXR for silicosis screening results in missed cases of silicosis and – in most scenarios – a relatively low number of participants are needed to be screened with HRCT to detect a single extra case of silicosis.

## Notation of prior abstract publication/presentation

Accepted for presentation by Ashwin Durairaj at the British Thoracic Society Winter meeting, London, United Kingdom, 27-29^th^ November 2024

## Data Availability

All data is available at https://github.com/pjhowlett/da_silic_cxr

## Acknowledgements

PH – Guarantor: full access to all the data in the study and takes responsibility for the integrity of the data and the accuracy of the data analysis, including and especially any adverse effects, Writing – Original draft preparation, Conceptualisation, Methodology, Project Administration, Data collection, Data Curation, Visualisation, Formal Analysis AD – Original draft preparation, Conceptualisation, Methodology, Project Administration, Data collection, Data Curation, Visualisation, Formal Analysis ML – Conceptualisation, Formal Analysis, Visualisation, Writing – Review & Editing JF – Conceptualisation, Project Administration, Supervision, Writing – Review & Editing

