## Supplement for "The diagnostic accuracy of chest Xray screening for silicosis: A systematic review, meta-analysis and modelling study"

**Supplementary Materials 1. Search strategy: Search strings used for Medline & Embase (A), Web of Science (B) and Scopus (C)**

| Search Number | Terms |
|---|---|
| 1 | exp Silicosis/ |
| 2 | silicos*.mp. |
| 3 | (Silicoses or silicosis).mp. |
| 4 | 1 or 2 or 3 |
| 5 | exp Tomography, X-Ray Computed/ or computer assisted tomography/ or thorax/ or computed tomography chest.mp. or thorax radiography/ or CT chest.mp. or HRCT.af. |
| 6 | exp X-Ray/ or (x adj ray*).af. or "radiogra*".af. |
| 7 | exp Autopsy/ |
| 8 | 5 or 6 or 7 |
| 9 | 4 and 8 |
| 10 | Clinical study/ or case-control study/ or retrospective study/ or cohort study/ or follow-up study/ or longitudinal study/ or prospective study/ or longitudinal study/ or cohort analysis/ or (Cohort adj (study or studies)).mp. or (follow up adj (study or studies)).mp. or (observational adj (study or studies)).mp. or (epidemiologic$ adj (study or studies)).mp. or (cross sectional adj (study or studies)).mp. or "comparative stud*".mp. or "comparison stud*".mp. or "Diagnostic accuracy of".mp. or "Diagnostic Value of".mp. |
| 11 | 9 and 10 |
| 12 | remove duplicates from 11 |
| 13 | limit 12 to English language |
| 14 | limit 13 to "remove preprint records" |

**B.**
ALL=(silicos?s) AND TS=(("X-ray" OR "xray" OR tomograph* OR (CT near/2 (scan* or screen*)) OR radiograph* OR HRCT)) AND (TS=(cohort stud* OR cohort analy* OR follow up stud* OR "follow-up" OR "follow up" OR "followup" OR observational stud* OR Longitudinal OR Retrospective OR cross sectional OR clinical stud* OR epidemiologic stud* OR prospective stud* OR prospectiv*OR comparative stud* OR comparison stud* OR “Diagnostic value of” OR “Diagnostic accuracy of”))

**C.**
TITLE-ABS-KEY("X-ray" OR "xray" OR "tomograph*" OR "CT scan" OR "CT scans" OR "CT screening" OR "radiograph*" OR HRCT) AND TITLE-ABS-KEY("cohort study" OR "cohort studies" OR "cohort analysis" OR "cohort analyses" OR "follow up study" OR "follow up studies" OR "follow-up" OR "followup" OR "observational study" OR "observational studies" OR "longitudinal" OR "retrospective" OR "cross sectional" OR "clinical study" OR "clinical studies" OR "epidemiologic study" OR "epidemiologic studies" OR "prospective study" OR "prospective studies" OR "prospectively" OR "comparative study" OR "comparative studies" OR "comparison study" OR "comparison studies" OR

“Diagnostic value of” OR “Diagnostic accuracy of”) AND ( LIMIT-TO ( LANGUAGE,"English" ) ) AND ( LIMIT-TO ( EXACTKEYWORD,"Silicosis" ) )

**Supplementary Materials 2. Data extraction template**

**General information**

- Study ID
- Title
- Journal
- Year of publication
- Country in which the study was conducted
- Conflicts of interest

**Characteristics of included studies**

Methods

- Study design
    - Retrospective cohort study
    - Prospective cohort study
    - Cross sectional study
    - Case control study
    - Diagnostic test accuracy study
    - Other study type
- Study start date
- Study end date

Participants

- Method of recruitment
    - Clinic patients
    - Occupational screening
    - Occupational health referral
    - Other
- Population description (employment, etc.)
- Mean age of population
- Mean years of estimated silica exposure
- Sample size
- Inclusion criteria
- Exclusion criteria
- Newly diagnosed or is this follow-up from pre-existing diagnosis?
- How did the study diagnose as positive for CXR?
- How did the study diagnose as positive for HRCT?
- How did the study diagnose as positive for autopsy?

**CXR vs (HR)CT**:

- Type of CT (normal CT, or HRCT)
- CXR positives
- CXR normal
- HRCT positives
- HRCT normal

2x2 Table of CXR vs (HR)CT

| | (HR)CT+ | (HR)CT- |
|---|---|---|
| CXR+ | | |
| CXR- | | |

CXR vs (HR)CT subgroup analysis

| | (HR)CT Cat 0 | (HR)CT Cat 1 | (HR)CT Cat 2 | (HR)CT Cat 3 |
|---|---|---|---|---|
| CXR ILO 0 | | | | |
| CXR ILO 1 | | | | |
| CXR ILO 2 | | | | |
| CXR ILO 3 | | | | |

**CXR vs autopsy:**

- CXR positives
- CXR normal
- Autopsy positives
- Autopsy negatives
- What is defined to be in each autopsy severity category?

2x2 Table: CXR vs Autopsy

| | Autopsy + | Autopsy - |
|---|---|---|
| CXR+ | | |
| CXR- | | |

CXR vs Autopsy Subgroup

| | Autopsy Cat 0 | Autopsy Cat 1 | Autopsy Cat 2 | Autopsy Cat 3 |
|---|---|---|---|---|
| CXR ILO 0 | | | | |
| CXR ILO 1 | | | | |
| CXR ILO 2 | | | | |
| CXR ILO 3 | | | | |

**Supplementary Materials 3. Implementation of the QUADAS-2 tool**

The QUADAS-2 tool, commonly used for diagnostic accuracy studies, was used to evaluate the risk of bias and applicability of included studies in our review. Below we outline its application across four domains for risk of bias, and provide examples from our included studies.

Domain 1: Patient selection

Could the selection of patients have introduced bias?

- Was a consecutive or random sample of patients enrolled?
- Was a case-control design avoided?
- Did the study avoid inappropriate exclusions?
-

Relevance: This evaluates whether patient selection could bias diagnostic accuracy. An unselected sample of silica-exposed workers is important to avoid inflating CXR sensitivity or specificity, which could occur if patients were pre-selected based on prior CXR results or clinical suspicion. Importantly, the issue is not whether the prevalence of silicosis is high or low, but whether there is a differential change, compared to the underlying population, of disease that is easier or harder to diagnose in the index test compared to the reference test.
Implementation: Studies were rated high risk if patients were pre-selected based on prior test results (e.g., abnormal CXR). Unclear risk was rated when sampling methodology was not described.
Example: Crawford et al., had high risk from selecting sandblasters to HRCT based on prior lung biopsies, where the decision to biopsy was prompted by “disparity between respiratory impairment and normal to mildly abnormal chest X-ray”. Hoy et al. had high risk from their selective enrolment, reliant in part on abnormal CXR, in order for secondary screening with HRCT.

Domain 2: Index test

Could the conduct or interpretation of the index test have introduced bias?

- Were the index test results interpreted without knowledge of the results of the reference standard?
- If a threshold was used, was it pre-specified?
-

Relevance: This evaluates if CXR interpretation was unbiased and standardised. Variability in reader expertise or lack of blinding could affect sensitivity/specificity.
Implementation: High risk was rated if CXR lacked a pre-specified threshold, or if information on the ILO readers was significantly insufficient (e.g., number of readers, reader training, or blinding to reference standard results). Unclear risk was rated when interpretation was described, but lacking regarding one aspect, e.g., blinding.
Separately, applicability concerns arose with non-standard thresholds (e.g., ILO >=0/1).
Example: Hoy et al., was rated high risk as details of the CXR evaluation were significantly insufficient, with unknown number of readers, their training, and methodology. Similarly, Crawford et al., provided no details on readers or blinding, and

lacked a pre-specified CXR positivity threshold. Grenier et al., was also rated high risk by not detailing the ILO threshold.

Domain 3: Reference standard

Could the reference standard, its conduct, or its interpretation have introduced bias?

- Is the reference standard likely to correctly classify the target condition?
- Were the reference standard results interpreted without knowledge of the results of the index test?
-

Relevance: This ensures the reference standards were unbiased by CXR results, important for making valid comparisons.
Implementation: High risk was rated if there was both unclear blinding to CXR and inadequate detailing of reference standard interpretation (e.g., number of readers, reader training). Unclear risk was rated when interpretation was described, but lacking regarding one aspect, e.g., blinding.
Example: Crawford et al., had unclear risk as there were no details on reference standard interpretation, including whether HRCT readers were blinded to CXR results. Separately in this domain, high applicability concerns arose for Meijer et al. from only using data on well-defined rounded opacities as their table overlapped results, and unclear applicability concerns for Takahashi et al. from an assumption their semi-quantitative HRCT scoring system at >=1 met the positive threshold for diagnosis.

Domain 4: Flow and timing

Could the patient flow have introduced bias?

- Was there an appropriate interval between index test(s) and reference standard?
- Did all patients receive a reference standard?
- Did patients receive the same reference standard?
- Were all patients included in the analysis?
-

Relevance: This evaluates if all patients underwent both tests consistently and if timing gaps could alter disease state, affecting diagnostic accuracy.
Implementation: High risk was rated for inconsistent reference standard application or significant intervals; although this was undefined, in practice we felt it was relevant to only one study (below). Unclear risk rated when timing was unreported.
Example: Hnizdo et al. had high risk due to an average 2.7-year gap between last CXR and autopsy, during which silicosis could progress, potentially reducing CXR sensitivity. Hoy et al., had high risk as 130 workers assessed in primary screening were excluded from the analysis due to not being determined as “high risk”, and thus did not have a HRCT.

**Supplementary Materials 4: Study-specific definitions of our reference standard severity categories**

The below definitions use language directly from the paper, unless stated otherwise.

| Talini 1995: HRCT severity categories | Definition |
|---|---|
| 0 | No definite nodules |
| 1 | Small number of nodules without disruption of vascular markings |
| 2 | Many definite nodules without confluence |
| 3 | Combines their grade 3 (confluence of nodules with disruption of vascular markings) and grade 4 (confluence of nodules extending over two or more slices) |

| Antao 2005: HRCT severity categories | Definition<br>(not from paper, inferred from referenced scales) |
|---|---|
| 0 | Absence of opacities/nodules |
| 1 | Small number of opacities/nodules not obliterating the vascular markings |
| 2 | Many opacities/nodules that slightly obliterate vascular markings, but without confluence. |
| 3 | Confluence of opacities/nodules, with the most severe blunting of the vascular markings, indicating severe disease. |

Note: The paper used a scale modified from two papers: Begin 1991 & Bergin 1986. The above is what we expect the definitions to represent, based on the two papers.

| Lopes 2008: HRCT severity categories | Definition |
|---|---|
| 0 | Absence of micronodules |
| 1 | Small number of micronodules without vascular blurring |
| 2 | Large number of micronodules, with or without vascular blurring, but with no confluence |
| 3 | Confluence of nodules <10mm, typically with pronounced vascular blurring |

| Bergin 1986: CT severity categories | Definition |
|---|---|
| 0 | No definite nodules |
| 1 | Small number of nodules with no disruption of vascular markings |
| 2 | Many nodules but without confluence |

| | |
|---|---|
| 3 | Combines their grade 3 (confluence of nodules, usually associated with disruption of vascular markings), and grade 4 (confluence of nodules extending over two or more slices, consistent with the diagnosis of progressive massive fibrosis) |

| Cowie 1993: CT severity categories | Definition |
|---|---|
| 0 | No nodules seen |
| 1 | Few nodules seen |
| 2 | Intermediate nodules seen |
| 3 | Innumerable nodules seen |

| Hnizdo 1993: Autopsy severity categories | Definition |
|---|---|
| 0 | No nodules, or an insignificant nodule |
| 1 | Few silicotic nodules |
| 2 | Moderate silicotic nodules |
| 3 | Large number of silicotic nodules |

| Corbett 1999: Autopsy severity categories | Definition |
|---|---|
| 0 | No discrete, palpable nodules in the two lungs |
| 1 | Occasional nodules (1-4), or early silicotic fibrosis (classified as “early grades of pathological silicosis”) |
| 2 | Few nodules (5-14) & moderate nodules (15-30) |
| 3 | Marked nodules (>30) |

**Supplementary Table 1.** Characteristics of studies which fit inclusion criteria. Reference numbers match those in the manuscript

Abbreviations: SD, standard deviation; CXR, chest Xray; HRCT, high-resolution computed tomography; CT, computed tomography; TP, true positive; FP, false positive; FN, false negative; TN, true negative; N/A, not available; ILO, International Labour Organization Classification; TB, tuberculosis; IOCERD, International Classification of High-resolution Computed Tomography for Occupational and Environmental Respiratory Diseases; MDT, Multidisciplinary team.

| **Study, country, design** | **Study period** | **Population Sample size (% male)** | **Sample size (% male)** | **Age; RCS exposure (Mean years (SD))** | **CXR classification, cut off and method** | **Reference standard classification and method** | **Result categories** | **Sensitivity (95% CI)** | **Specificity (95% CI)** |
|---|---|---|---|---|---|---|---|---|---|
| HRCT | | | | | | | | | |
| Bégin 1991, Canada, Cross-sectional[17] | N/A | Workers' Compensation Board referrals; exposure in mine/foundries | 49 (NA) | 57.1 (1.4); 29.2 (1.7) | ILO ≥ 1/0; 4 readers (2 NIOSH-B readers). Blinded, independent. Mean score. | Scale with same principles as ILO. Both CT and HRCT read. Method as CXR | TP 12<br>FP 1<br>FN 17<br>TN 19 | 0.41 (0.23, 0.61) | 0.95 (0.75, 1.00) |
| Grenier 1991, France, Cross-sectional[18] | 1986 - 1989 | Subgroup of study of various chronic diffuse interstitial lung diseases | 11 (NA) | N/A; N/A | ILO (unclear cut-off) ; 3 “experienced” radiologists. Blinded, independent. Median score | Unknown. Method as CXR | TP 2<br>FP 0<br>FN 9<br>TN 0 | 0.18 (0.02, 0.52) | 1.00 (0.03, 1.00) |
| Talini 1995, Italy, Cross-sectional[19] | N/A | Screened exposed workers in mine, glass, building and pottery industries | 27 (96) | 57 (8.8); N/A | ILO ≥ 1/0; 2 “experienced” readers. Unclear blinding. Independent. MDT consensus | Bergin (1986) scale. Method as CXR | TP 14<br>FP 5<br>FN 6<br>TN 2 | 0.70 (0.46, 0.88) | 0.29 (0.04, 0.71) |
| Antao 2005, Brazil, Cross-sectional[20] | N/A | Current stone carvers; worked at least 1 year | 41 (97.5) | 36.2 (8.7); N/A | ILO ≥ 1/0; 3 trained A-readers. Blinded, independent. Median score | Modified scale from Begin (1991). Method as CXR | TP 19<br>FP 3<br>FN 3<br>TN 16 | 0.86 (0.65, 0.97) | 0.84 (0.60, 0.97) |
| Murgia 2007, Italy, Cross-sectional[21] | 2004 – 2005 | Artisan gold and silversmiths | 100 (97) | 42.2 (9.9); 18.1(9.8) | ILO ≥ 1/0; 2 “experienced” radiologists. Blinded, independent. Disagreement then 3rd radiologist | Scale from Begin (1991). Method as CXR | TP 10<br>FP 0<br>FN 13<br>TN 77 | 0.43 (0.23, 0.66) | 1.00 (0.95, 1.00) |
| Sun 2008, China, Cross-sectional[22] | N/A | Male workers engaged in sandcasting | 90 (100) | 51 (4.8); 15.7 (2.8) | GBZ70-2002* (ILO ≥ 1/0); 5 "experienced" readers. Blinded. Independent. Consensus if ≥3/5 agreement, else median. | Own scale, principles from GBZ70-2002. Method as CXR | TP 30<br>FP 0<br>FN 5<br>TN 55 | 0.86 (0.70, 0.95) | 1.00 (0.94, 1.00) |
| Lopes 2008, Brazil, Cross-sectional[23] | N/A | Mainly sandblasters (45.5%), stone cutters (34.1%). Non-smokers, no prior TB | 44 (93) | 48.4; 16.1 | ILO ≥ 1/0; 3 “trained” readers. Unclear blinding, independent. Median score | Own scale, ILO principles. MDT consensus of 4 radiologists | TP 40<br>FP 0<br>FN 4<br>TN 0 | 0.91 (0.78, 0.97) | 1.00 (0.03, 1.00) |

| | | | | | | | | | |
|---|---|---|---|---|---|---|---|---|---|
| Meijer 2011, Netherlands, Cross-sectional[24] | 2002-2002 | 180 construction workers with different stages on CXR from a prior 1998 study were re-invited in 2002 | 77 (100) | 49.6; 21.2 | ILO ≥ 1/1; 2 B readers. Blinded, independent. Consensus with 3rd B reader as needed | German Federal Republic classification 3 B-readers. Blinded, independent. MDT Consensus | TP 3<br>FP 0<br>FN 9<br>TN 65 | 0.25 (0.06, 0.57) | 1.00 (0.94, 1.00) |
| Tamura 2015, Japan, Cross-sectional[15] | N/A | Patients from clinic for suspected pneumoconiosis with 28 controls | 74 (100) | N/A; 31 | ILO ≥ 1/0; 3 B readers. Unclear blinding. Independent. Median results | ICOERD. 3 (different) radiologists. Unclear blinding. Independent. Median results | TP 20<br>FP 3<br>FN 1<br>TN 50 | 0.95 (0.76, 1.00) | 0.94 (0.84, 0.99) |
| Berk 2016, Turkey, Cross-sectional[25] | N/A | All dental technicians working in Sivas, Turkey | 32 (94) | 31 (9); 14 (9) | ILO ≥ 1/0; 3 readers (1 B reader). Blinded, independent. MDT consensus | ICOERD. Method as CXR | TP 8<br>FP 0<br>FN 14<br>TN 10 | 0.36 (0.17, 0.59) | 1.00 (0.69, 1.00) |
| Takahashi 2018, Japan, Cross-sectional[26] | 2014 - 2015 | Silicosis cases from tertiary hospital | 33 (91) | 74; 37.8 | Japan Pneumoconiosis Law Classification^ (≥ 1/0); 2 "experienced" radiologists. Blinded, independent. MDT consensus | Modified ICOERD. Method as CXR | TP 25<br>FP 1<br>FN 4<br>TN 3 | 0.86 (0.68, 0.96) | 0.75 (0.19, 0.99) |
| Şener 2019, Turkey, Cross-sectional[27] | 2012-2014 | Foundry workers, welders, or miners with pneumoconiosis and CXR and HRCT | 83 (100) | 44.5 (11.5); 16.5 | ILO ≥ 1/0; 2 B-reader physicians. Blinded. MDT consensus | ICOERD. 3 radiologists. MDT consensus | TP 72<br>FP 0<br>FN 7<br>TN 4 | 0.91 (0.83, 0.96) | 1.00 (0.40, 1.00) |
| León-Jiménez 2020, Spain, Retrospective cohort[28] | 2009 - 2018 | Male factory/installation workers with artificial stone silicosis | 106 (100) | 36.2 (7); 12 (4.3) | ILO ≥ 1/0; 3 "trained" readers. Unclear blinding. MDT consensus | ICOERD. Method as CXR | TP 87<br>FP 0<br>FN 19<br>TN 0 | 0.82 (0.73, 0.89) | 1.00 (0.03, 1.00) |
| Hoy 2023, Australia, Cross-sectional[29] | 2019 - 2021 | Secondary screening of artificial stone benchtop workers | 396 | Median 38 (IQR 30,47); Median 10 (IQR 5, 15) | ILO ≥ 1/0; unknown reader experience and method | Own scale. Method unclear | TP 74<br>FP 40<br>FN 36<br>TN 246 | 0.67 (0.58, 0.76) | 0.86 (0.81, 0.90) |
| Crawford 2024, United States, Retrospective cohort[30] | 1988-1995 | Workers sandblasting with oil-field drilling equipment in West Texas | 42§ (100) | 35.6 (10.4); 7.6 (4.8) | Custom rating system of 4 scores^ (≥ 1). Unknown readers, blinding, independence or consensus method | Bergin (1986) scale. Unknown methods | TP 21<br>FP 0<br>FN 3<br>TN 18 | 0.88 (0.68, 0.97) | 1.00 (0.81, 1.00) |
| CT | | | | | | | | | |
| Bergin 1986, Canada, Cross-sectional[31] | 1984 - 1985 | Hard rock mining or sandblasting workers with exposure to silica | 23 | 58.6; 19.1 | ILO ≥ 1/0; 1 experienced radiologist. Blinded, independent. | Own scale, ILO principles. 2 readers, read twice. Median score | TP 17<br>FP 0<br>FN 1<br>TN 5 | 0.94 (0.73, 1.00) | 0.50 (0.21, 0.79) |
| Bégin 1987, Canada, Cross-sectional[17] | N/A | Granite or foundry workers in Quebec | 58 | 59 (2); 30 | ILO ≥ 1/0; 3 readers. Unknown experience and method | Own scale, ILO principles. 3 readers. Unknown experience $e and method | TP 46<br>FP 6<br>FN 0<br>TN 6 | 1.00 (0.92, 1.00) | 1.00 (0.48, 1.00) |

| | | | | | | | | | |
|---|---|---|---|---|---|---|---|---|---|
| Cowie 1993, South Africa, Cross-sectional[34] | N/A | Subset from larger cohort of 1197 older gold miners (ref?) | 70 (100) | 49.7 (5.9); 29.6 (7.8) | ILO ≥ /1; 2 readers. Unclear experience. Blinded, independent. Unclear consensus | Own scale, ILO principles. >1 reader however number and method unclear | TP 46<br>FP 9<br>FN 2<br>TN 13 | 0.96 (0.86,0.99) | 0.59 (0.36, 0.79) |
| Autopsy | | | | | | | | | |
| Hnizdo 1993, South Africa, Retrospective cohort[8] | 1968 - 1971 | Subset of cohort of 2,260 White South African gold miners | 557 | 50.6 (2.48); 27.1 (6.6) | ILO ≥ 1/0 (reader 2 only); 3 experienced readers. Blinded, independent. Separately reporting | 5-point scale of fibrosis/nodules. 5 pathologists; 1 per autopsy | TP 163<br>FP 25<br>FN 163<br>TN 206 | 0.50 (0.44, 0.56) | 0.89 (0.84, 0.93) |
| Corbett 1999, South Africa, Cross-sectional[33] | 1996 - 1997 | Male gold miners who died between Jan 1996 - July 1997 | 241 (100) | N/A, N/A | ILO ≥ 1/0; 2 "experienced" readers. Blinded, independent, MDT consensus | 5-point scale of fibrosis/nodules. 5 pathologists; 1 per autopsy | TP 40<br>FP 12<br>FN 40<br>TN 149 | 0.50 (0.39, 0.61) | 0.93 (0.87, 0.96) |

^ ILO equivalent. * Not possible to compare to ILO system. § Paper does not explicitly state number of CT scans performed; number extrapolated from % of abnormal scans reported (24, 57%)

**Supplementary Table 2.** Sensitivity analysis: Missed cases and number needed to screen in a silica-exposed population. Calculated using the metaregression model with exposure of prevalent ILO >2/1 silicosis. We present the number of missed cases per 1000 persons and number needed to screen to identify 1 extra case of silicosis in three scenarios of low silicosis prevalence (5% or 50 cases per 1000), as defined by CXR ILO >1/0, medium silicosis prevalence (15% or 150 cases per 1000) and high silicosis prevalence (30% or 300 cases per 1000), and under the assumptions of fixed and relative sensitivity of CXR, compared to HRCT. For the assumption of relative sensitivity, we have further assumed either less common severe disease (20% of cases are ≥ ILO 2/1) and more common severe disease (40% of cases are ≥ ILO 2/1).

| | | Low silicosis prevalence (5%) | | Medium silicosis prevalence (15%) | | High silicosis prevalence (30%) | |
|---|---|---|---|---|---|---|---|
| | | Missed cases per 1000 exposed | NNS to detect 1 extra case | Missed cases per 1000 exposed | NNS to detect 1 extra case | Missed cases per 1000 exposed | NNS to detect 1 extra case |
| Fixed CXR sensitivity | | 16 | 62 | 47 | 21 | 95 | 11 |
| Relative CXR sensitivity | Less severe disease (20% > ILO 2/1) | 69 | 14 | 189 | 5 | 334 | 3 |
| | More severe disease (40% > ILO 2/1) | 66 | 15 | 167 | 6 | 260 | 4 |

**Supplementary Figure 1.** Forest plots of sensitivity and specificity from meta-analysis of chest X-ray (CXR), at an ILO ≥1/0 cut-off, for the diagnosis of silicosis. The figure describes the sensitivity (left) and specificity (right) of CXR compared to computed tomography (CT).

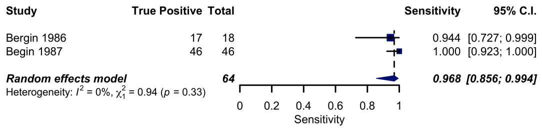

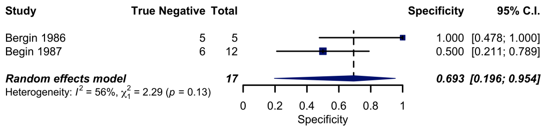

**Supplementary Figure 2.** Assessment of methodological quality of included twenty studies using the Quality Assessment of Diagnostic Accuracy Studies-2 tool.

- **A.** Risk of bias and applicability summary: review authors' judgements about each domain for each included study.
- **B.** Risk of bias and applicability plot: review authors' judgements about each domain presented as percentages across included studies

A

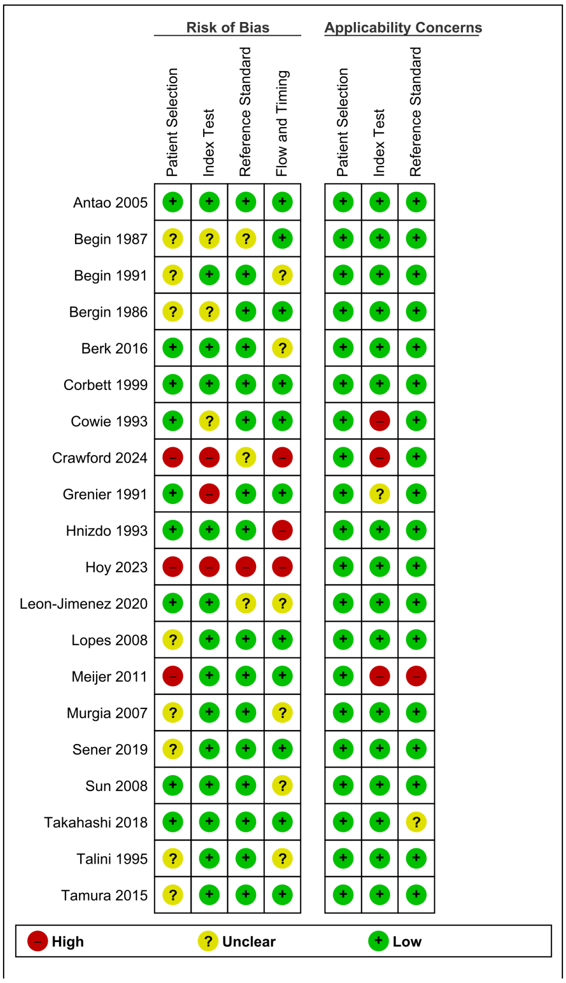

B

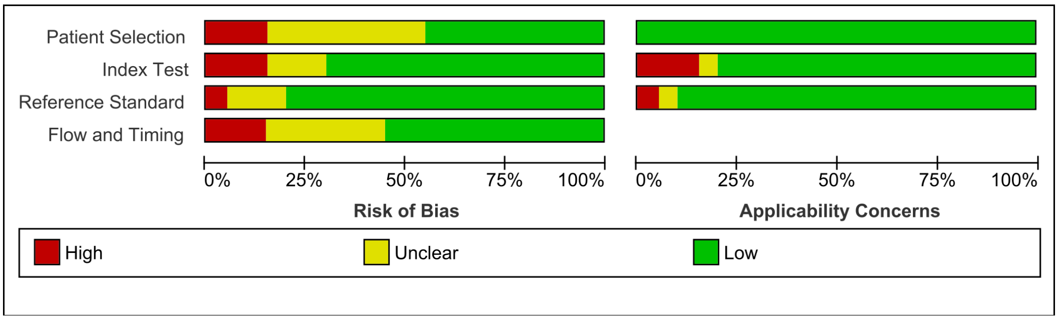

**Supplementary Figure 3.** Sensitivity analysis: Forest plots of sensitivity and specificity from meta-analysis of chest X-ray (CXR) in the diagnosis of silicosis including additional ILO and non-ILO cut-offs.

**A.** Describes the sensitivity (left) and specificity (right) of CXR compared to high-resolution computed tomography (HRCT).

**B.** Describes the sensitivity (left) and specificity (right) of CXR compared to computed tomography (CT).

A

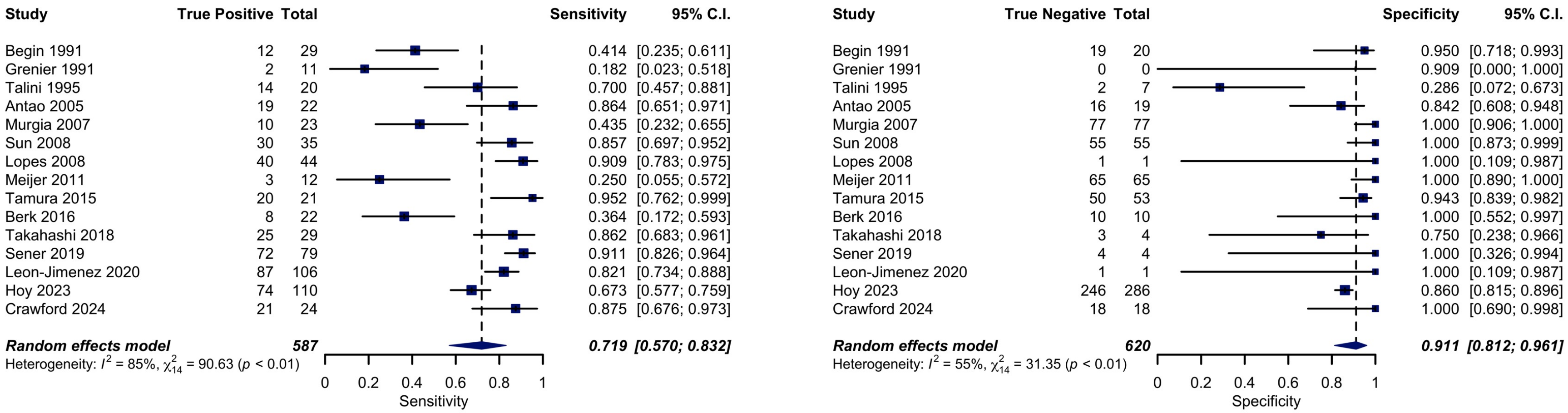

B

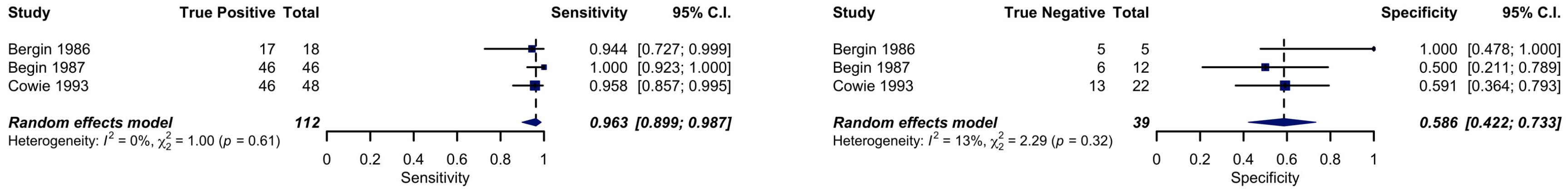

**Supplementary Figure 4.** Sensitivity analysis: Forest plots of sensitivity (left) and specificity (right) from meta-analysis of chest X-ray (CXR) in the diagnosis of silicosis compared to high-resolution computed tomography (HRCT), including removal of studies with a high/unclear risk of patient selection or index test bias.

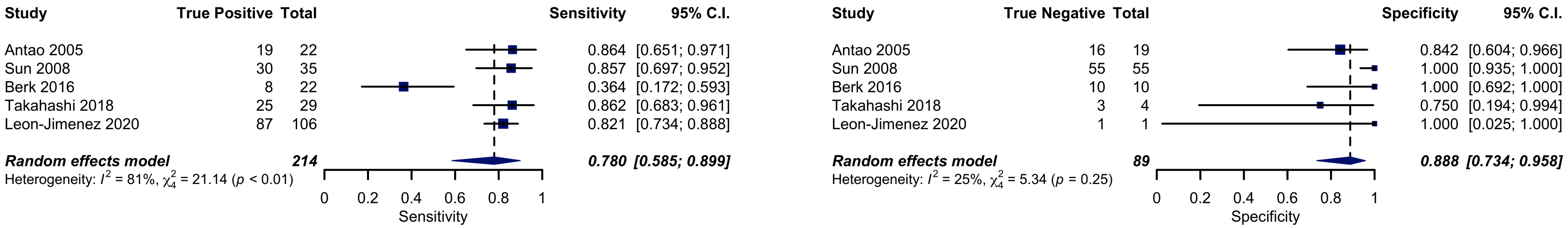

**Supplementary Figure 5.** Forest plots of sensitivity (left) and specificity (right) from with the reference test at higher grades of radiology reference test

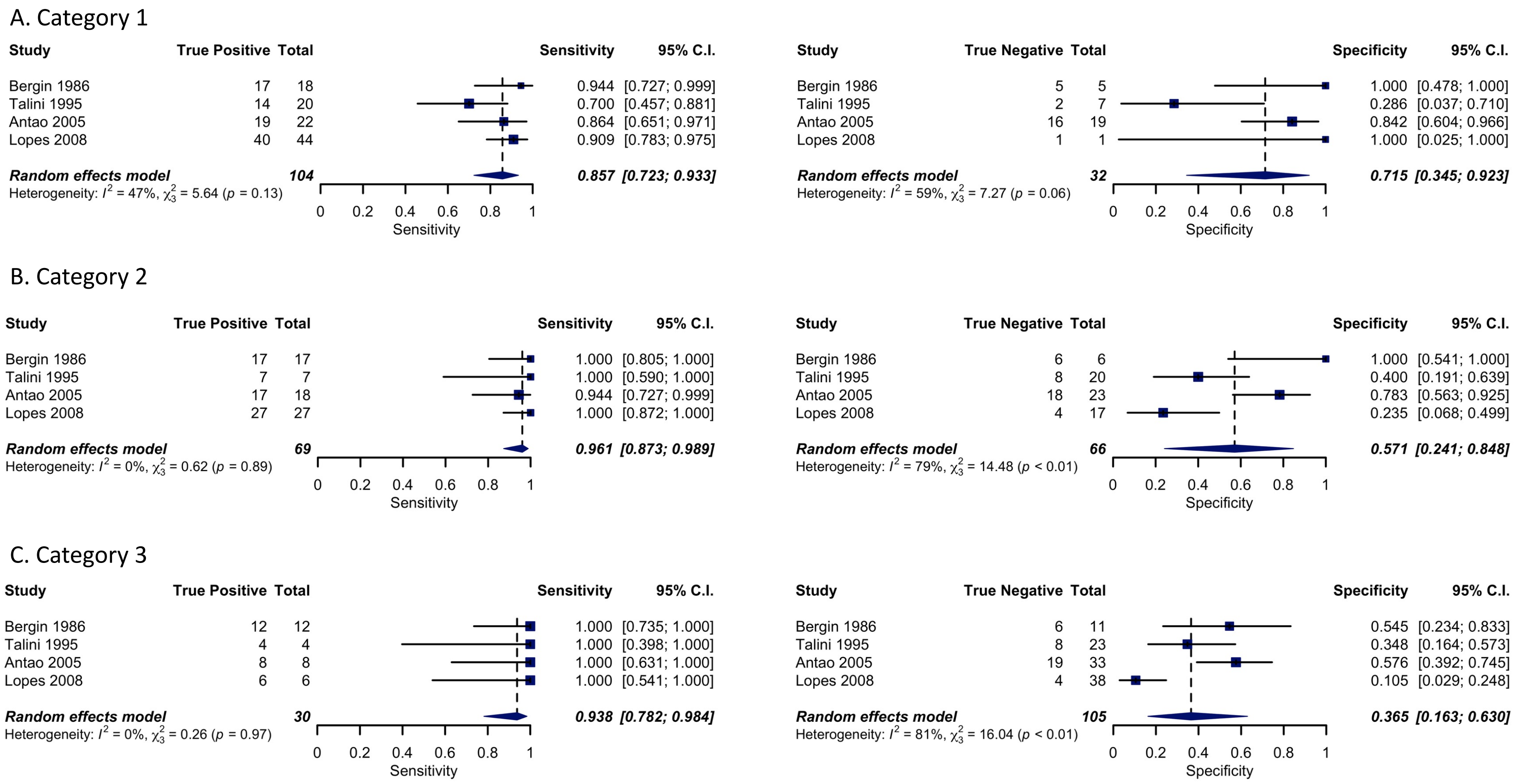

**Supplementary Figure 6.** Forest plots of sensitivity (left) and specificity (right) from with the reference test at higher grades of autopsy reference test.

A. Category 1

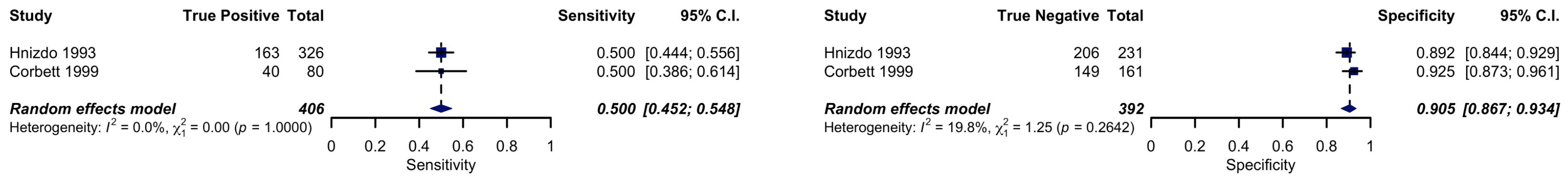

B. Category 2

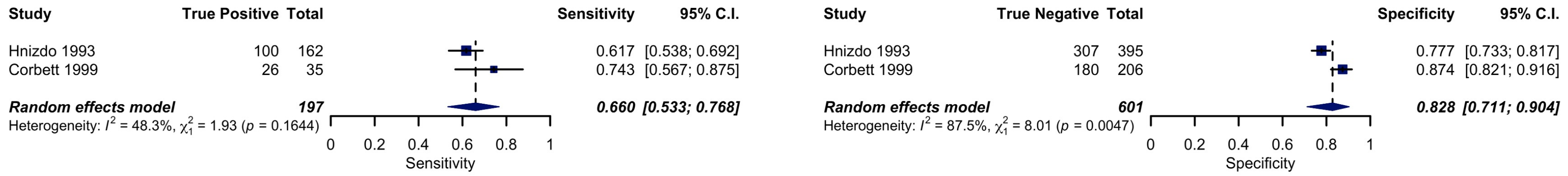

C. Category 3

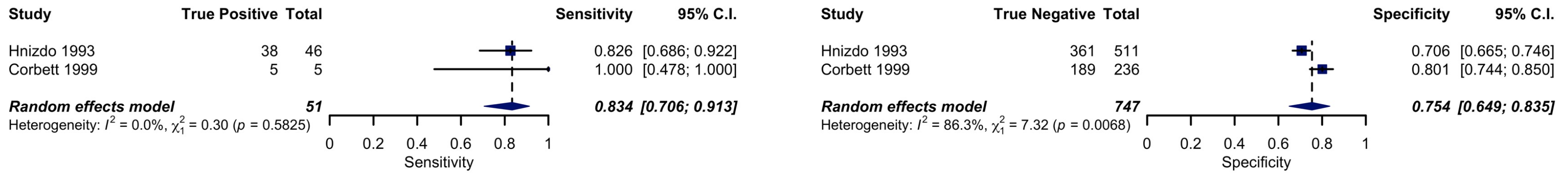

**Supplementary Figure 7.**

Missed silicosis cases. Presents the estimated number of cases missed per 1000 exposed persons across range of silicosis prevalences (defined by ILO > 1/0 from 0 to 40%. Three scenarios of fixed and relative sensitivity of CXR, both compared to refrence of HRCT are presented. Fixed sensitivity is labelled "Fixed". For the assumption of relative sensitivity, we have assumed either less common severe disease (20% of cases are > ILO 2/1), labelled "Relative: 20% severe" and more common severe disease (40% of cases are > ILO 2/1), labelled "Relative: 40% severe"

A. Calculates the relative sensitivity using the sensitivity analysis metaregression and the exposure of the ratio of CXR ILO ≥2/1 silicosis relative to CXR ILO ≥ 1/0 silicosis

B. Calculates the relative sensitivity using the primary analysis metaregression and the exposure of prevalent CXR ILO ≥2/1 silicosis in the study population.

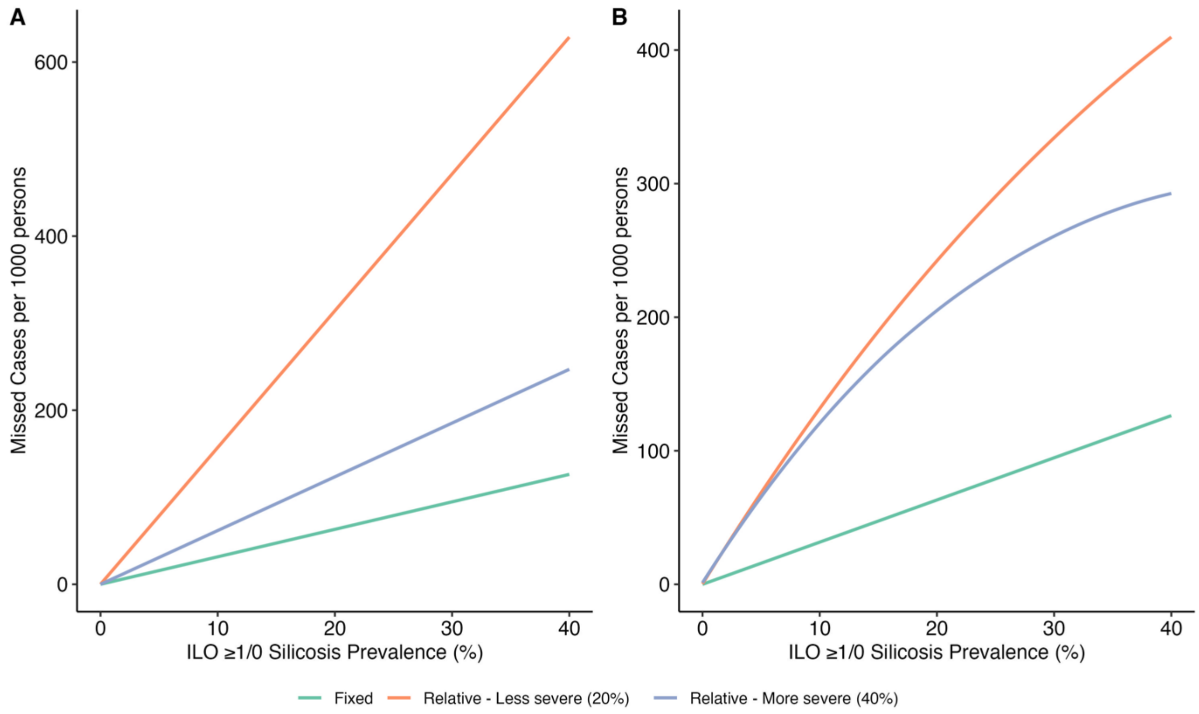

**Supplementary Figure 8**. Sensitivity analysis: Relationship between severity of silicosis, silica exposure and sensitivity of chest Xray (CXR). Mixed-effects linear meta-regression model showing the association between the sensitivity of CXR at ILO category ≥1 and the prevalence of CXR ILO >2/1 (our sensitivity analysis) in 7 studies. Each circle represents a study, with the size proportional to the study's weight in the meta-regression. The circle is coloured depending on reference standard (Red = autopsy, Blue = computed tomography and Green = high resolution computed-tomography). The solid line represents the predicted sensitivity based on the proportion of severe cases. The shaded areas represent the 95% confidence interval. The $R^2$ = 94% and heterogeneity is $I^2$ = 50%.

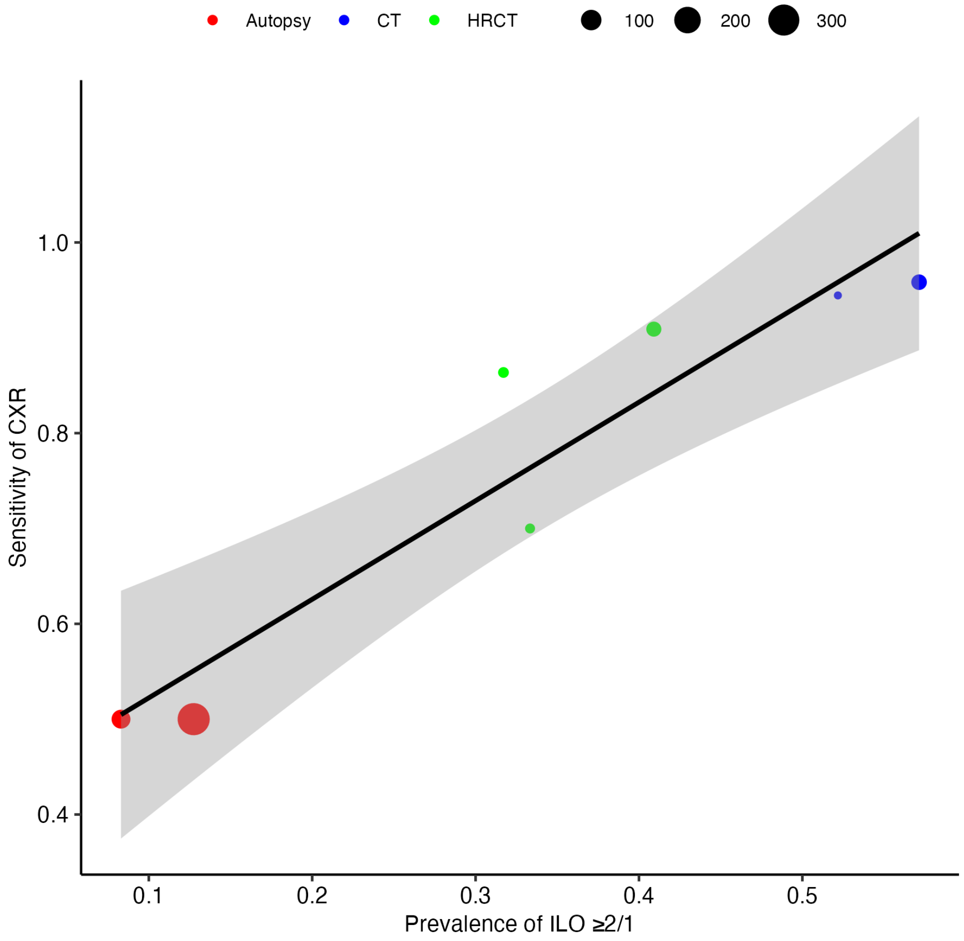
